# Updated Results of 3,050 Non-melanoma Skin Cancer (NMSC) Lesions in 1725 Patients Treated with High Resolution Dermal Ultrasound-Guided Superficial Radiotherapy, A Multi-institutional Study

**DOI:** 10.1101/2023.04.18.23288760

**Authors:** Mairead Moloney, Peter Kaczmarksi, Songzhu Zheng, Ariana Malik, Daniel Ladd, Donna Serure, Lio Yu

## Abstract

**Background:** Image-guided superficial radiation therapy (IGSRT) using a high resolution dermal ultrasound, is becoming an attractive non-surgical curative treatment option for nonmelanoma skin cancer (NMSC). We previously reported IGSRT treatment results from a multi-institutional study of 1616 patients with 2917 NMSC lesions (Yu *et al*. Oncol Ther 2021, https://doi.org/10.1007/s40487-021-00138-4) showing excellent local control (LC) of 99.3% with mean follow-up of 69.8 weeks (16.06 months).

**Methods:** This abstract analyzes an additional 93 patients with 133 lesions, updates previous findings with longer follow-up, using Kaplan-Meier statistics, and performs subgroup analysis. A total of 1709 patients with 3,050 Stage 0, I, and II NMSC lesions treated from 2017 to 2020 were retrospectively analyzed.

**Results:** Lesions received a median of 20 fractions of 50, 70, or 100 kilovoltage(kV) IGSRT using image guidance. Average follow-up was 25.06 months (108.9 weeks) with a maximum follow up of 65.6 months (285.0 weeks) for the entire cohort. 68 patients expired from unrelated causes with no-evidence of disease (NED) at last follow-up prior to death, thus Disease-Free-Survival (DFS) was 100%. Overall, 3,027 of 3,050 lesions achieved an absolute LC of 99.2%. Overall absolute LC for BCC, SCC, and SCCis was 99.0%, 99.2%, and 99.8%, respectively. No additional late complications were found to date as of January 2022.

**Discussion:** These updated results demonstrate that IGSRT continues to achieve a high level of LC with low complication rates. IGSRT should be considered an attractive first-line option for the non-surgical treatment of NMSC.

## INTRODUCTION

Non-melanoma skin cancer (NMSC) is the most common cancer in the United States (US) (1). The overwhelming majority of NMSC are comprised of basal cell carcinoma (BCC) and squamous cell carcinoma (SCC), which account for 99% of NMSC. Since NMSC are not reported to national cancer registries, the most current estimate of NMSC occurrences are from 2012, where it was estimated that there was 5.43 million NMSC lesions in the United States (US) population and 3.32 million patients treated for NMSC (2). The incidence of NMSC is expected to be increasing by two to three percent yearly (3). In 2023, this translates to 6.75 to 7.52 million lesions in 4.13 to 4.60 million individuals. Additionally, individuals often have more than one NMSC lesion and the incidence of subsequent lesions increases after initial lesion diagnosis (3).

NMSC is considered nonfatal and curable due to its slow growth, low recurrence, and rare metastasis (4). It is typically treated via surgical modalities, including Mohs Micrographic Surgery (MMS). However, NMSC occurs on the head and neck in 70-80% of cases, and surgery can leave scars and cosmetic defects (5). There are numerous non-surgical treatment modalities that exist with moderate control rates. Image-Guided Superficial Radiotherapy (IGSRT) which incorporates a high resolution dermal ultrasound has emerged recently as a modality that has excellent control rates comparable to MMS and is an attractive non-surgical treatment modality for the treatment of NMSC (5),(6).

From March 2016 to January 2020 a retrospective chart review of 1616 patients from seven out-patient dermatology practices with 2917 early-stage non-melanoma skin cancer (NMSC) lesions treated with image-guided superficial radiotherapy (IGSRT) was conducted. The study’s objective was to assess the efficacy and safety of IGSRT in treating a large number of patients with NMSC lesions. In June 2021, the results were published, which showed an excellent local control (LC) of 99.3% with mean follow-up of 69.8 weeks (16.06 months) (5). These results have continued to allow physicians to offer IGSRT to their patients as a treatment for their early-stage NMSC lesions.

This purpose of this analysis is to add an additional 93 patients with 133 lesions from another out-patient dermatology practice and update the previous results with longer follow-up and provide a subgroup analysis.

## METHODS

The details regarding patient selection, treatment guidelines, study endpoints, and statistical analyses were discussed in the previous publication, and remain consistent for this updated analysis (Yu *et al*. Oncol Ther 2021, https://doi.org/10.1007/s40487-021-00138-4)(5). Statistical methods were augmented with Kaplan-Meier analysis.

### PATIENTS

A retrospective chart review of 1709 patients, including 93 additional patients treated with IGSRT for NMSC between March 2016 and January 2020 at 8 outpatient dermatology practices (7 previous dermatology practices, 1 additional dermatology practice) across the USA were analyzed. Lesion characteristics and treatment data at the time of treatment were collected retrospectively during the chart review.

Prior to treatment with IGSRT, lesion diagnosis and staging were confirmed via biopsy performed by a dermatologist at each practice.

The study cohort comprised of 756 female and 953 male patients (mean age 74.04 years [SD ± 10.64]) with 3,050 lesions treated from years 2016 to 2020. Each lesion was considered an independent cancer lesion and all lesions included in this analysis were stage Tis, T1, or T2. There was no clinical evidence of regional lymph node or distant disease (N0 and M0) at presentation, based on the American Joint Committee on Cancer (AJCC) 8^th^ Ed.

Cancer Staging Manual (7). The AJCC 8^th^ Ed. staging is specific to cutaneous SCC of the head and neck, but for consistency these same criteria were applied to all BCC, SCC, and SCCIS lesions in this study.

Initially, data and follow-up intervals were extracted manually from written and electronic medical records. Subsequent updates to patient data and follow-up were accessed electronically via algorithmic analysis provided by a healthcare data company (Sympto Health, Inc.).

The authors adhered to the principles established in the Federal Policy for the Protection of Human Subjects, referred to as the “Common Rule,” as well as the pertinent sections of the Helsinki Declaration and its amendments. The study protocol was reviewed by an IRB committee (WIRB-Copernicus Group) and received IRB exemption. The data have been de-identified for use in this study.

### TREATMENT

Treatment characteristics are summarized descriptively in **Table 1**. All lesions received a median of 20 fractions (range 13-30 fractions) of 50, 70, 100, or mixed (ie. 50/70, 70/100, 50/70/100) kilovoltage (kV) IGSRT. Ultrasound imaging allowed for energy selection before and during treatment. The median dose per fraction was 258.72 cGy (mean 259.24 cGy [SD +/- 10.42]). The median total dose received was 5184.3 cGy (mean 5215.95 cGy [SD +/- 222.25]). Median treatment duration was 6.71 weeks (mean 6.94 weeks [SD +/- 1.53]). Median time dose fraction (TDF) was 90 (mean 90.82 [SD +/- 4.76]). During IGSRT treatment, patients were evaluated at each session clinically and with ultrasound imaging and treatment dosing adjustments were made if necessary.

**Table 1.** Treatment characteristics for tumor treatment with Image-Guided Superficial Radiation Therapy (IGSRT).

| IGSRT Treatment Characteristics |  |  |
| --- | --- | --- |
| **Total Dose Received (cGy)** | Mean | 5215.95 (SD +/- 222.25) |
|  | Median | 5184.3 (IQR: 5096, 5336) |
|  | Range | 3715.95 - 7363.68 |
| **Number of Fractions** | Mean | 20.13 (SD +/- 0.72) |
|  | Median | 20 (IQR: 20, 20) |
|  | Range | 13 - 30 |
| **Dose Per Fraction (cGy)** | Mean | 259.24 (SD +/- 10.42) |
|  | Median | 258.72 (IQR: 254.40, 265.47) |
|  | Range | 14.56 - 401.76 |
| **Treatment Duration (weeks)** | Mean | 6.94 (SD +/- 1.53) |
|  | Median | 6.71 (IQR: 6.14, 7.43) |
|  | Range | 4 – 22.43 |
| **Time Dose Fractionation (TDF)** | Mean | 90.82 (SD +/- 4.76) |
|  | Median | 90 (IQR: 87.53, 94) |
|  | Range | 75.86 - 138 |

### FOLLOW-UP

Once treatment ended patients were evaluated 2-6 weeks after treatment completion and every 1-6 months thereafter.

### STUDY ENDPOINTS

This study’s main outcome measure was lesion recurrence assessed by local control (LC) and Kaplan-Meier (KM) LC analysis with follow-up through January 2022. LC was assessed at follow-up visits clinically by dermatology providers. IGSRT’s safety was assessed by Radiation Treatment Oncology Group (RTOG) toxicity, which was prospectively documented in the charts routinely after every 5-fractions. RTOG data was extracted and documented as the highest RTOG grade from the entire treatment course. Certain study locations did not maintain a procedure to record RTOG toxicity grades for IGSRT patients in the initial study year(s), accounting for the missing values on this measure of safety/toxicity.

### STATISTICAL METHODS

Treatment information, patient demographics, tumor characteristics, and RTOG toxicity were summarized descriptively. Treatment information included total dose received (cGy), number of fractions, dose per fraction (cGy), energy (kV), treatment duration, and time dose fractionation (TDF)^A^. Duration of follow-up was defined as the date of last follow-up minus treatment completion date plus one day and then converted to both weeks and months.^B^ Patient demographics included total patients, age at first treatment, gender, follow-up interval, and deaths. Tumor characteristics included number of lesions, lesion histopathology, lesion size, lesion recurrences, and lesion location. Lesion histopathology separated by tumor stage and lesion histopathology separated by energy treatment were also provided. Acute toxicities were graded with RTOG (Radiation Treatment Oncology Group) toxicity scoring (8).

Summary statistics for continuous outcomes included the number of observations (n), mean, standard deviation, median, interquartile range, and range. Categorical outcomes were summarized with frequency and percentages. Missing data were not included. SAS® Studio was used for all analyses and verification with R Studio.

All patients were included in the intent-to-treat analysis. Overall LC and KM LC were reported. KM LC was reported based on lesion histology was compared using log-rank tests.

Subgroup analyses of patients with ≥12-month follow-up, ≥24-month follow-up, and breakdown by histology (BCC, SCC, SCCIS) were performed. Number of lesions, lesions recurred, absolute local control, overall 5-year KM LC, and 5-year KM LC by histology subtype were reported.

For this updated report, the results are based on all the information received by January 14^th^, 2022.

## RESULTS

### PATIENT DEMOGRAPHICS AND TUMOR CHARACTERISTICS

**Table 2** details patient demographics. Median age at first treatment was 74.45 years (mean 74.04 years [SD +/- 10.64 years]). There was slight predominance of male patients at 55.8% (953 patients) with 756 female patients (44.2%). Median follow-up was 25.38 months (mean 25.06 months [SD +/- 17.10 months]).

**Table 2.** Patient demographics at time of lesion treatment with Image-Guided Superficial Radiation Therapy (IGSRT).

| Patient Demographics |  |  |
| --- | --- | --- |
| Total Patients |  | 1709 |
| Age at 1st Treatment (Years) | Mean | 74.04 (SD +/- 10.64) |
|  | Median | 74.45 (IQR: 68.05, 81.59) |
|  | Range | 31.80 - 104.52 |
| Gender | Female | 756 |
|  | Male | 953 |
| Follow-up Interval (Months) | Mean | 25.06 (SD +/- 17.10) |
|  | Median | 25.38 (IQR: 9.45, 38.43) |
|  | Range | 0.03 - 65.59 |
| Death* (Number of Patients) |  | 68 |

\*Death from other causes

Tumor characteristics are described in **Table 3**. All lesions were stage Tis, T1 or T2 with BCC representing 47.9%, followed by SCC (30.3%) and SCCIS (21.2%). Median lesion size was 1.0 cm (mean 1.23 cm [SD +/- 0.65 cm]), showing that these are substantial lesions. Lesion histopathology separated by lesion stage is depicted in **Table 4**. Lesion histopathology separated by treatment energy is depicted in **Table 5**. Lesion histopathology separated by lesion location is depicted in **Table 6**.

**Table 3.** Tumor characteristics at time of lesion treatment with Image-Guided Superficial Radiation Therapy (IGSRT).

| Tumor Characteristics |  |  |
| --- | --- | --- |
| **Total Lesions** |  | 3050 |
| **Histopathology** | BCC | 1460 |
|  | SCC | 924 |
|  | SCCIS | 648 |
|  | BCC / SCC | 11 |
|  | BCC / SCCIS | 1 |
|  | SCC / SCCIS | 2 |
|  | BCC / SCC / SCCIS | 0 |
|  | No Data Recorded | 4 |
| **Lesion Size (cm)\*\*** | Mean | 1.23 (SD +/- 0.65) |
|  | Median | 1 (IQR: 1, 1.5) |
|  | Range | 0.05 - 5.5* |
| **Recurrences**<br>**(Number of**<br>**Lesions)** | BCC | 15 |
|  | SCC | 7 |
|  | SCCIS | 1 |
|  | BCC / SCC | 0 |
|  | BCC / SCCIS | 0 |
|  | SCC / SCCIS | 0 |
|  | BCC / SCC / SCCIS | 0 |
|  | Total Recurrences | 23 |

\*SCCIS (Squamous Cell Carcinoma In-Situ) all with full thickness atypia
\*\*28 lesions with no size recorded

**Table 4.**
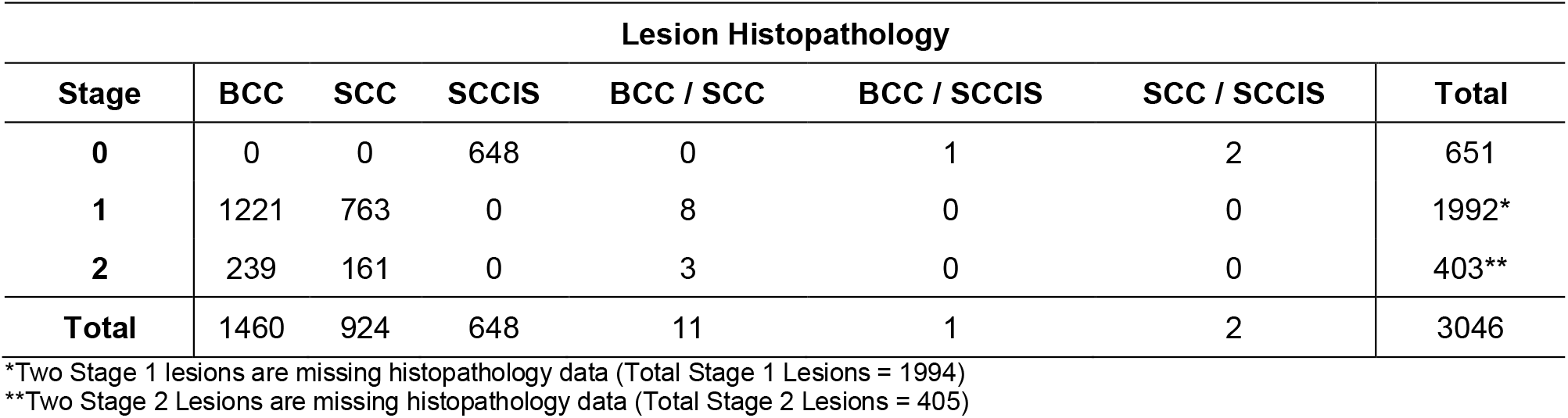
Lesions separated by histopathology and tumor stage.

**Table 5.**
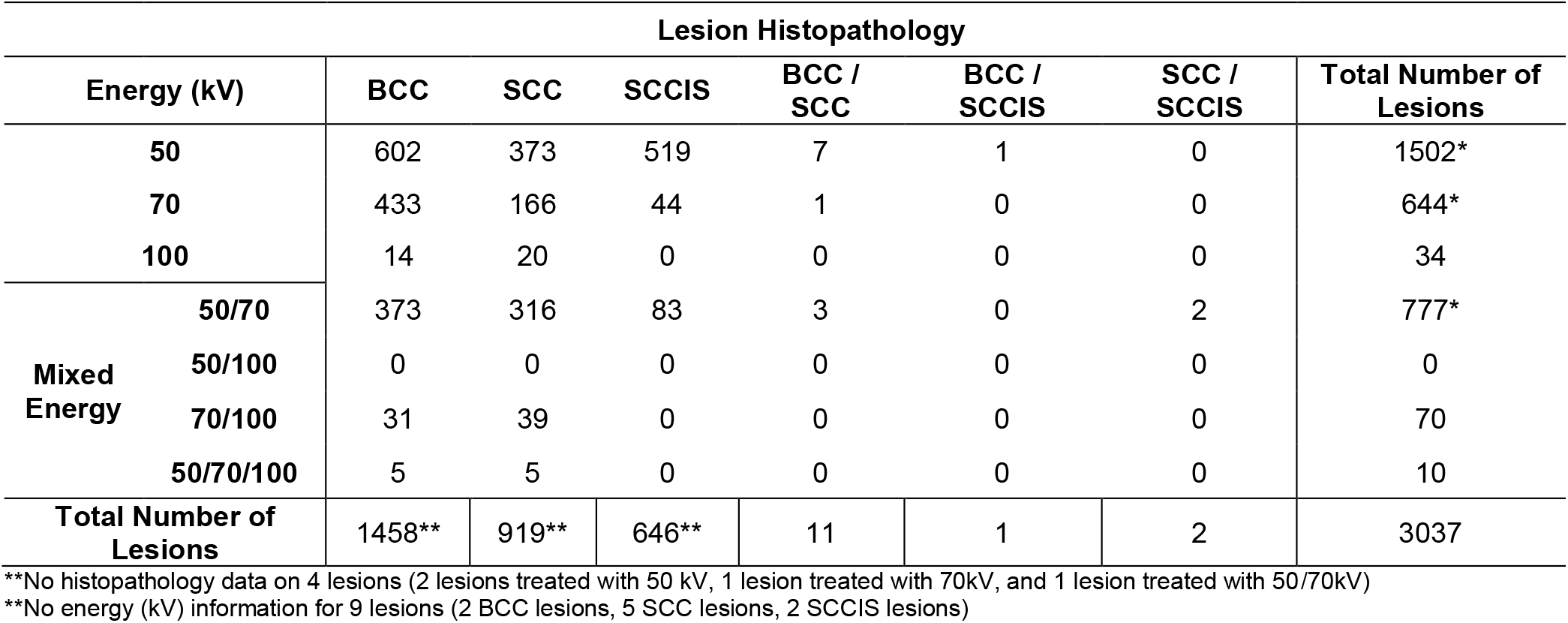
Lesions separated by histopathology and energy (kV) treatment.

**Table 6.** Lesions separated by histopathology and lesion location.

| Site | BCC,<br>N=1457* | SCC,<br>N=923* | SCCIS,<br>N=647* | BCC/SCC,<br>N=11 | BCC/SCCIS,<br>N=1 | SCC/SCCIS,<br>N=2 | Total,<br>N=3041** |
| --- | --- | --- | --- | --- | --- | --- | --- |
| **Head and Neck**<br>**(H&N)** | 1065 | 551 | 405 | 8 | 1 | 2 | 2032 |
| *H&N sublocation* |  |  |  |  |  |  |  |
| **Ear** | 124 | 95 | 55 | 2 | 0 | 0 | 276 |
| **Scalp** | 53 | 84 | 59 | 0 | 0 | 1 | 197 |
| **Forehead** | 112 | 79 | 86 | 0 | 0 | 0 | 277 |
| **Temple** | 41 | 20 | 12 | 1 | 0 | 0 | 74 |
| **Forehead/Temple** | 0 | 0 | 1 | 0 | 0 | 0 | 1 |
| **Eyebrow** | 5 | 2 | 4 | 0 | 0 | 0 | 11 |
| **Eyelid & Canthus** | 24 | 2 | 1 | 0 | 0 | 0 | 27 |
| **Nose** | 373 | 81 | 52 | 3 | 0 | 0 | 509* |
| **Cheek** | 204 | 122 | 105 | 2 | 1 | 1 | 435* |
| **Cutaneous Lip** | 51 | 13 | 7 | 0 | 0 | 0 | 71* |
| **Mucosal Lip** | 5 | 13 | 4 | 0 | 0 | 0 | 22 |
| **Chin** | 7 | 0 | 0 | 0 | 0 | 0 | 7 |
| **Jawline** | 2 | 3 | 0 | 0 | 0 | 0 | 5 |
| **Chin/Jawline** | 1 | 0 | 0 | 0 | 0 | 0 | 1 |
| **Neck** | 63 | 36 | 19 | 0 | 0 | 0 | 118 |
| **Other** | 0 | 1 | 0 | 0 | 0 | 0 | 1 |
| **Extremities** | 173 | 310 | 197 | 1 | 0 | 0 | 681* |
| *Extremities sublocation* |  |  |  |  |  |  |  |
| **Hand** | 11 | 78 | 57 | 0 | 0 | 0 | 146 |
| **Other** | 162 | 232 | 140 | 1 | 0 | 0 | 535 |
| **Shoulder** | 61 | 15 | 11 | 0 | 0 | 0 | 87 |
| **Trunk** | 158 | 46 | 34 | 2 | 0 | 0 | 240 |
| *Trunk sublocation* |  |  |  |  |  |  |  |
| **Chest** | 46 | 26 | 13 | 0 | 0 | 0 | 85 |
| **Back** | 101 | 20 | 21 | 1 | 0 | 0 | 143 |
| **Other** | 11 | 0 | 0 | 1 | 0 | 0 | 12 |
| **Penis** | 0 | 1 | 0 | 0 | 0 | 0 | 1 |

\*5 lesion locations not recorded (3 BCC, 1 SCC, 1 SCCIS)
\*\*4 lesions with no recorded histopathology (1 extremity lesion [not a hand], 1 nose lesion, 1 cheek lesion, 1 cutaneous lip lesion)

As of January 2022, 1,641 of 1,709 patients were alive. 68 patients expired from unrelated causes with no-evidence of disease (NED) at last follow-up prior to death, thus Disease-Free-Survival (DFS) was 100%.

### OUTCOMES

#### ABSOLUTE LOCAL CONTROL (LC)

At a mean follow-up of 25.06 months, 3,027 of 3,050 lesions achieved local control with 23 recurrences (15 BCC, 7 SCC, 1 SCCIS), resulting in an overall absolute LC of 99.2%. Absolute local control for BCC, SCC and SCCIS were 99.0%, 99.2%, and 99.8%, respectively.

#### KAPLAN MEIER LOCAL CONTROL (KM LC) at ≥5 Years

Overall 5-year KM LC was 98.81% and unchanged at maximum follow-up of 65.56 months (**Figure 1**). KM LC for BCC was 98.17% at 5-years (60 months). KM LC for SCC was 99.01% at 5 years (60 months) (**Figure 2**). KM LC for SCCIS was 99.71% at 5-years (60 months). Comparison of KM LC between histologic subtypes was not statistically significant using log-rank tests (p=0.0630, alpha=0.05).

**Figure 1.**
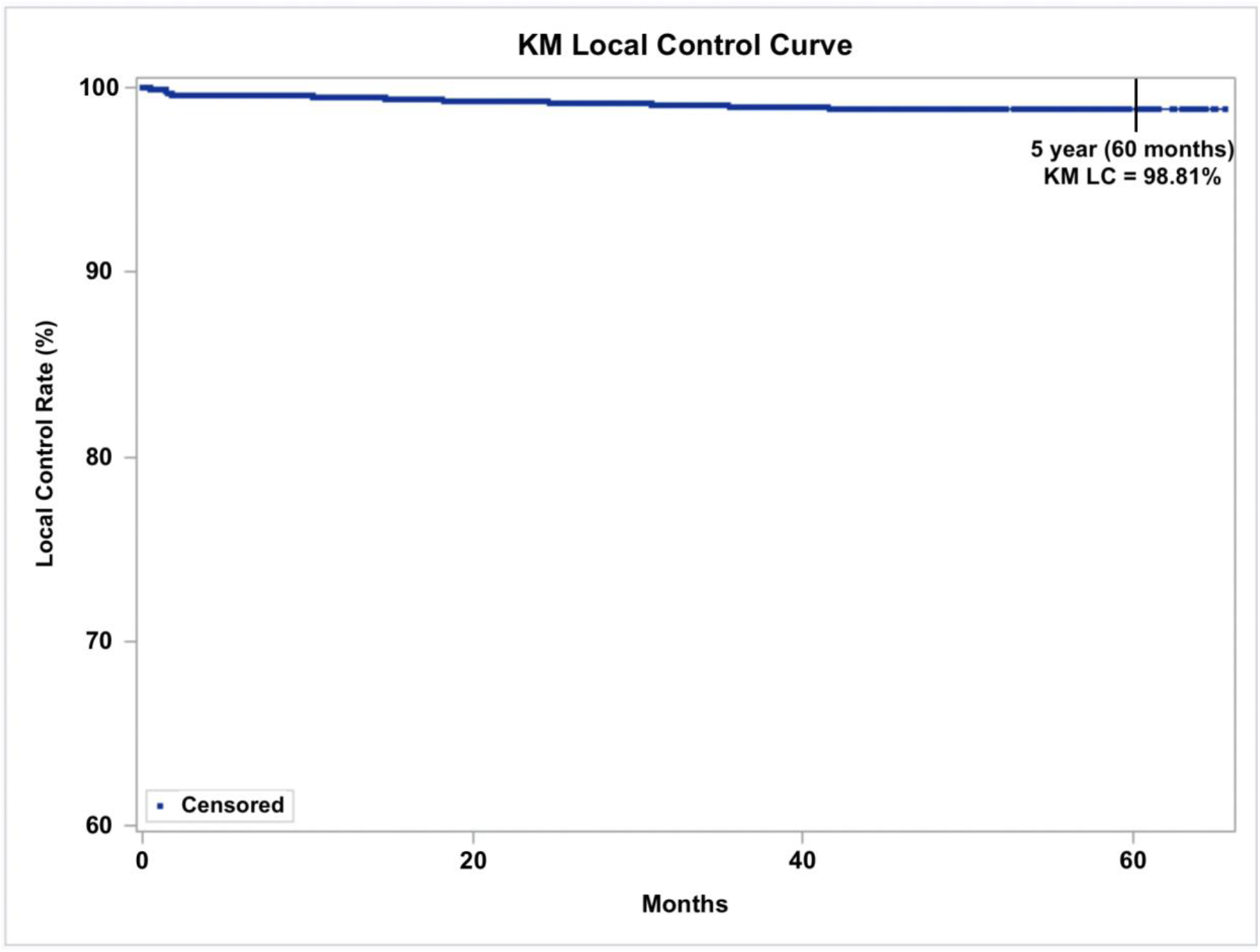
Kaplan-Meier Local control (KM LC) for all 3,050 lesions treated with IGSRT (Image-Guided Superficial Radiation Therapy). Dots represent censored events. (Note: Y-axis starts at 60%).

**Figure 2.**
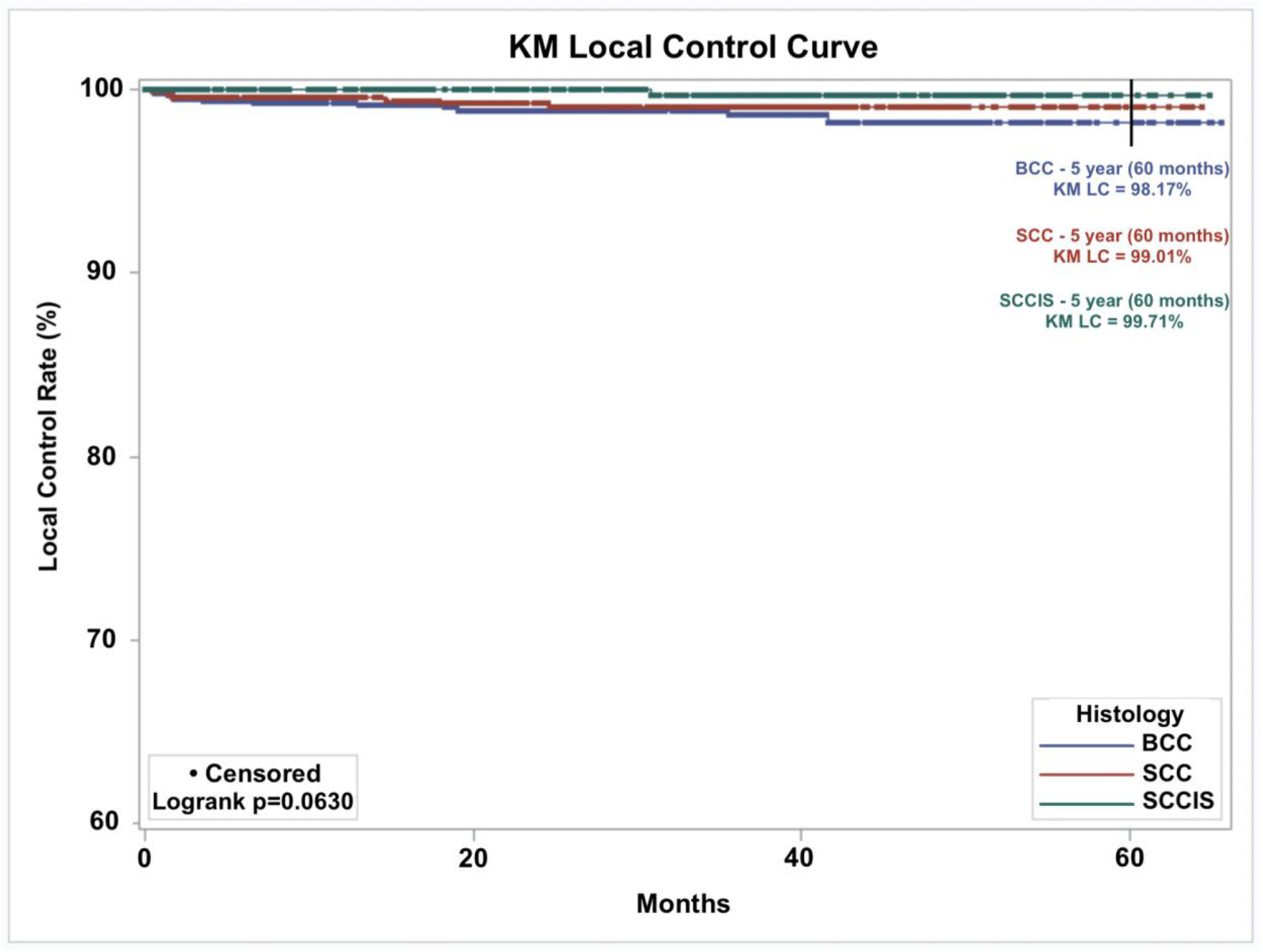
Kaplan-Meier Local control (KM LC) by histologic subtype of 3,032 lesions (Basal Cell Carcinoma (BCC) = 1,460 lesions, Squamous Cell Carcinoma (SCC) = 924 lesions, SCC In-Situ (SCCIS) = 648 lesions) treated with IGSRT (Image-Guided Superficial Radiation Therapy). Dots represent censored events. (Note: Y-axis starts at 60%).

### TREATMENT TOLERANCE

All lesions had minimal or mild toxicity (RTOG 0, 1, 2) with only 20 lesions having severe or significant toxicity (RTOG 3, 4) based on the RTOG toxicity scoring (8) (**Table 7**). No additional late complications were found to date as of January 2022.

**Table 7.** Acute toxicities RTOG (Radiation Treatment Oncology Group) grades for lesions treated with IGSRT (Image-Guided Superficial Radiation Therapy) (8).

| RTOG | Number of Lesions (n = 2,310)* |
| --- | --- |
| 0 | 1 |
| 1 | 1818 |
| 2 | 471 |
| 3 | 16 |
| 4 | 4 |

\*Certain study locations did not maintain a procedure to record RTOG toxicity grades for IGSRT patients in the initial study year(s), accounting for the missing values on this measure of safety.

### SUBGROUP ANALYSIS

Subgroup analysis of lesions with follow-up greater than or equal to 12 months and 24 months is summarized in **Table 8** and **Table 9**.

**Table 8.** Subgroup analysis comparing absolute local control (LC) and overall 5-year Kaplan Meier local control (KM LC) for all lesions, lesions with follow-up ≥ 12-months, and lesions with follow-up ≥ 24-months.

| Subgroup Analysis |  |  |  |
| --- | --- | --- | --- |
|  | All Lesions | >= 12-Month Follow-up | >= 24-Month Follow-up |
| Number of Lesions | 3050 | 2174 | 1615 |
| Lesions Recurred | 23 | 9 | 4 |
| Absolute Local Control (LC) | 99.2% | 99.6% | 99.8% |
| Overall 5-Year Kaplan Meier Local Control (KM LC) | 98.81% | 99.32% | 99.57% |

**Table 9.** Subgroup analysis comparing 5-year Kaplan Meier local control (KM LC) separated by histopathologic subtype (BCC, SCC, SCCIS only) for all lesions, lesions with follow-up ≥ 12-months, and lesions with follow-up\ ≥ 24-months.

| Subgroup Analysis |  |  |  |
| --- | --- | --- | --- |
|  | All Lesions | >= 12-Month Follow-up | >= 24-Month Follow-up |
| Number of Lesions | 3032 | 2163* | 1608* |
| 5-Year KM LC - BCC | 98.17% | 98.96% | 99.30% |
| 5-Year KM LC - SCC | 99.01% | 99.48% | 99.80% |
| 5-Year KM LC - SCCIS | 99.71% | 99.71% | 99.71% |

\*Lesions with mixed histology or no histology recorded were not included.

#### Subgroup with ≥12-Month Follow-up

A total of 2174 lesions had a follow-up time of greater than or equal to 12 months, representing 71.3% of all lesions. A total of 9 lesions recurred. Resulting in an overall absolute LC of 99.6% and overall KM LC was 99.32% at 5-years (60 months). KM LC for BCC lesions was 98.96%, for SCC lesions was 99.48%, and for SCCIS lesions 99.71% at 5-years (60 months) (**Figure 3**). Comparison of KM LC between histologic subtypes was not statistically significant (p=0.5776, alpha=0.05).

**Figure 3.**
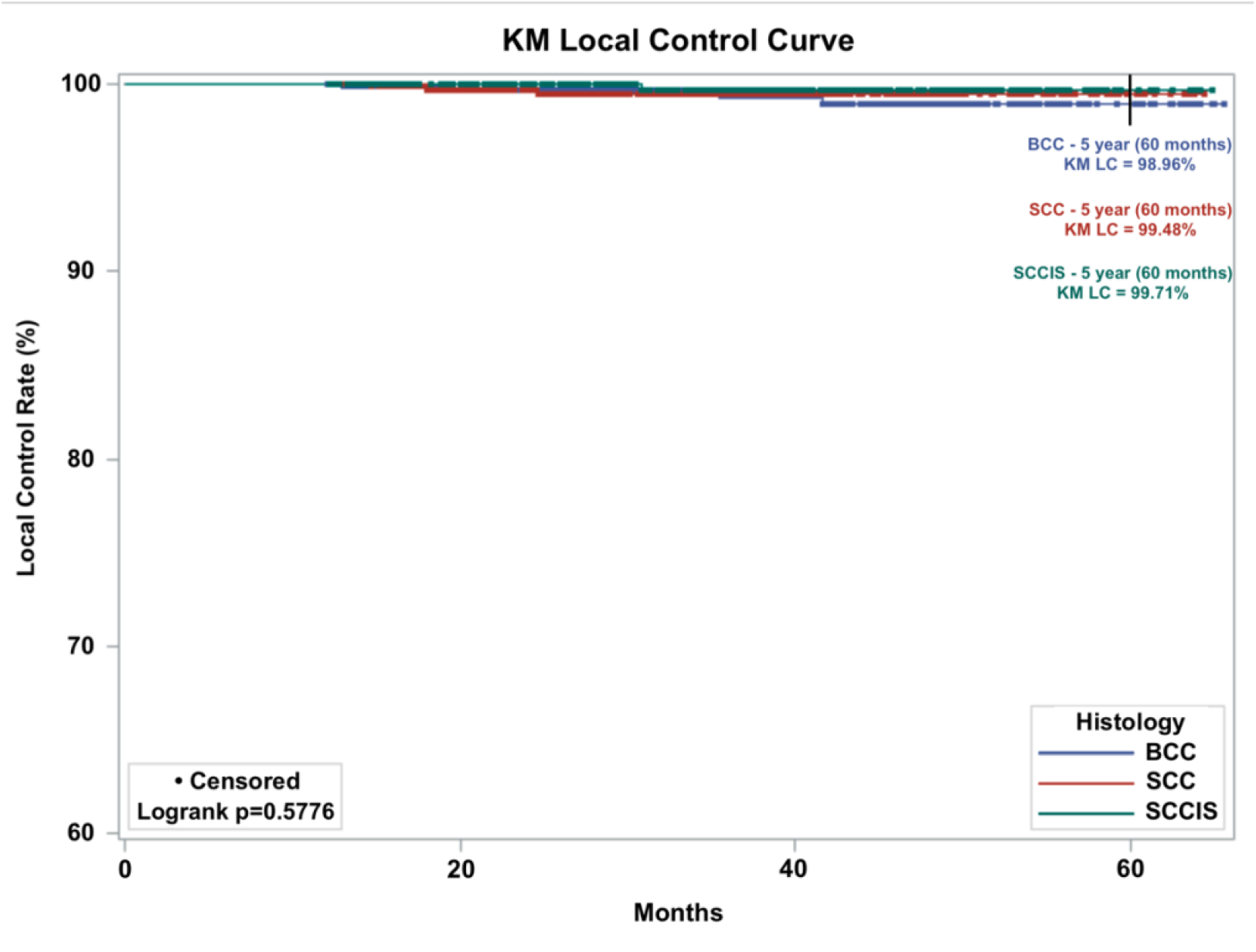
Kaplan-Meier Local control (KM LC) by histologic subtype of 2,174 lesions treated with IGSRT (Image-Guided Superficial Radiation Therapy) with a follow-up of greater than or equal to 12 months. Dots represent censored events. (Note: Y-axis starts at 60%).

#### Subgroup with ≥24-Month Follow-up

A total of 1615 lesions had a follow-up time of greater than or equal to 24 months, representing 53.0% of all lesions. A total of 4 lesions recurred. Resulting in an overall absolute LC of 99.8% and overall KM LC was 99.57% at 5-years (60 months). KM LC for BCC lesions was 99.30%, for SCC lesions was 99.80%, and for SCCIS lesions 99.71% at 5-years (60 months) (**Figure 4**). Comparison of KM LC between histologic subtypes was not statistically significant (p=0.9301, alpha=0.05).

**Figure 4.**
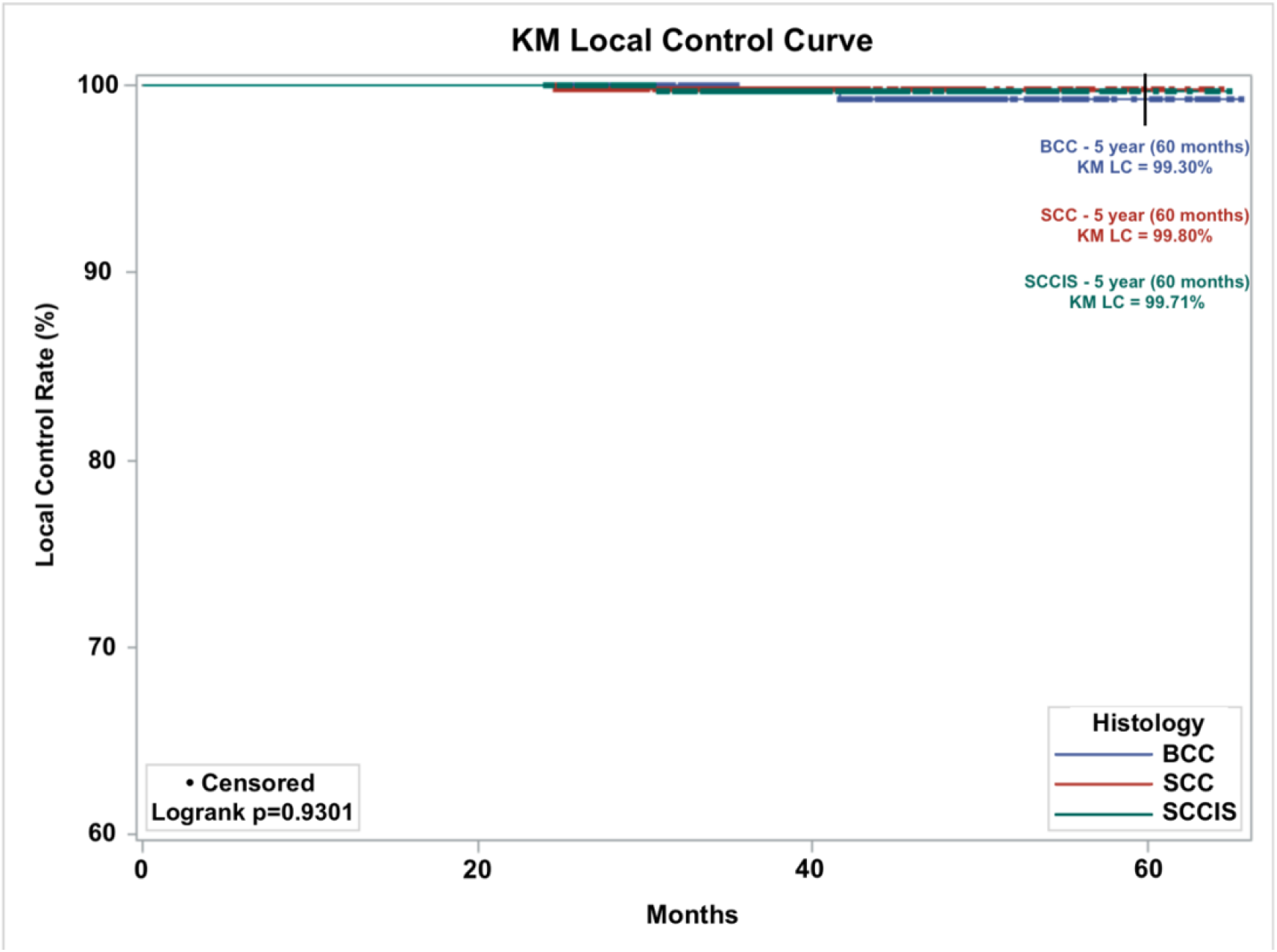
Kaplan-Meier Local control (KM LC) by histologic subtype of 1,615 lesions treated with IGSRT (Image-Guided Superficial Radiation Therapy) with a follow-up of greater than or equal to 24 months. Dots represent censored events. (Note: Y-axis starts at 60%).

## DISCUSSION

This analysis updates the results of the largest multi-institutional analyses of the utilization of image-guided superficial radiation therapy (IGSRT) for the curative treatment of early-stage NSMC. Our updated results support the initial findings and this analysis with longer follow-up has a similar absolute LC rate to previously published results.

Absolute LC rate of 99.2% (mean follow-up 25.06 months) versus the previously reported local control rate of 99.3% (mean follow-up 16.06 months), show that the absolute LC remains substantively unchanged with additional patients and longer follow-up (5).

Currently there are few modern studies on the use of IGSRT or even non-image guided SRT for the treatment of early-stage NMSC. An extensive literature search conducted by the American Society for Radiation Oncology (ASTRO) from 1988 to 2018 showed that there were limited randomized controlled trials (RCT) on radiotherapy (9). Additionally, our own PubMed search revealed that there are no published studies looking at the use of IGSRT for the curative treatment of NMSC, besides the previous analysis that this manuscript updates and an additional two manuscripts published by the authors (4), (5), (6). One of the more recently published analyses by the authors that utilized IGSRT for the curative treatment of NMSC lesions demonstrated an overall absolute LC of 99.7% and a 5-year KM LC of 99.41% for 1899 NMSC lesions in 1243 patients (6). The study by Tran et al. utilized a different cohort of patients having similar histology and stages of lesions showing the results were comparable to our current analysis which updated the 2021 study demonstrating that IGSRT yields consistently excellent results.

Our updated results have a mean follow-up of greater than 2 years with approximately 3/4 of the lesions analyzed having a follow-up of greater than or equal to 1 year (71.3%) and approximately 1/2 of the lesions analyzed having a follow-up of greater than or equal to 2 years (53.0%). In the previously published manuscript, we compared our results to a study by Cognetta et al. from 2012 that utilized non-image guided SRT and is one of the more modern studies using SRT to treat NMSC with curative intent. However, at that time only 55% of our patients had a follow-up greater than or equal to 1 year. The study by Cognetta et al. reported a 2-year recurrence rate for BCC and SCC lesions at 2.0% and 1.8% for BCC and SCC, respectively (10). Thus, the 2-year absolute LC would be 98.0% and 98.2% for BCC and SCC, respectively. Our results continue to be consistent and exceed those reported by Cognetta et al.

The likelihood of local recurrence for early-stage NMSC is small after two years. A majority (70-80%) of SCC are thought to recur within the first 2 years after primary tumor treatment with 95% of SCC recurrences occurring within 5 years (11). Whereas for BCC, approximately 50% of recurrences occur within the first 2 years after treatment of the primary tumor with 80% of BCC recurrences occurring with 5 years (12). Although some reports suggests that BCC recurrences primarily occur within the first 4-12 months (11). Our results corroborate with the latter.

Additionally, we previously compared our preliminary results to another SRT study by Hernández-Machin et al. (13). Hernández-Machin et al. reported 5-year absolute LC rates of 94.4% for BCC lesions and 92.7% for SCC lesions. With these results, the authors recommended SRT as a first line treatment option for NMSC. Our estimated 5-year KM LC remains higher at 98.81%. This is consistent with our belief that image guidance is most likely responsible for the improved outcomes.

Our improved absolute LC results over those of other studies that utilized SRT alone or external beam radiation therapy (XRT) without image guidance, is statistically significant. Two recent studies, a metanalysis and a logistic regression analysis, compared IGSRT studies to large well run SRT/XRT studies without image guidance. Both analyses found that the LC rates for the IGSRT studies were statistically superior to the non-image-guided SRT/XRT studies (14), (15). The image-guided component of SRT utilizes a 22 megahertz (MHz) dermal ultrasound with color doppler to visualize the superficial depth of the skin. This image-guided component allows the physician to visualize the lesion prior to, during, and after treatment. During treatment, the physician can make any necessary adjustments to the treatment regimen.

Our large patient sample from multiple institutions support that’s this office-based technology, IGSRT, is feasible, effective, safe and easily tolerable in an out-patient setting. Almost all lesions had minimal or mild toxicity scores (RTOG 0, 1, 2). Our updated results support that IGSRT, with longer follow-up, continue to offer a high local control rate felt to be comparable to the best surgical techniques (14) (15) and continues to be an effective non-invasive, non-surgical treatment modality for NMSC with its major advantages of avoiding the pitfalls of surgery clinically, cosmetically, physiologically and psychologically. These updated results validate our prior statement that IGSRT should be a primary non-surgical option presented to patients for treatment of their early-stage NMSC lesions.

## Data Availability

Deidentified data are available on request from Dr. Lio Yu at. Data will be available for request with publication for period of at least 1 year.

## Footnotes

A The IGSRT treatment protocol has evolved over the past 5-6 years, specifying time dose fractionation (TDF) number/dose/fractionation based on ultrasound depth and tumor type. This protocol recommends a fractionation dose range of 245-279 cGy for 20 fractions 3-4 times a week to achieve a therapeutic biological dose range of 90-99 or greater TDF number using 50, 70, or 100 kV energy. Higher doses per fraction and/or more fractions were recommended for larger, deeper, and high-risk lesions (5).

B If follow-up date was unavailable, treatment completion date was used as the last follow-up date. If treatment completion date was unavailable, treatment start date was used as the treatment completion date and last-follow-up date. Days were converted to months using the approximation of 30.417 days per month and days were converted to weeks using 7 days per week.

